# The Prognostic Value of CanAssist Breast in Patients ≤50 Years: A Retrospective Study

**DOI:** 10.64898/2026.08.04.26359653

**Authors:** Manvi Sunder, Tejal D. Durgekar, Shyamili Goutham, Badada Ananthamurthy Savitha, Payal Shrivastava, Naveen Krishnamoorthy, Deepti K Shivashimpi, Koushik Anand S J, Anjali Raghuram, Manjiri M. Bakre

**Affiliations:** OncoStem Diagnostics Private Limited, 4, Raja Ram Mohan Roy Road, Aanand Towers, 2nd Floor, Bangalore, Karnataka 560027, India

**Keywords:** CanAssist Breast, young patients, early-stage breast cancer, chemotherapy, hormone-receptor positive, prognostication

## Abstract

**Background:** Patients aged ≤50 years with early-stage HR+/HER2-breast cancer are considered to have an aggressive disease biology and are treated with chemotherapy. However, a subset may still experience favourable outcomes without chemotherapy. Commercially available prognostic tests help guide such treatment decisions, but most are developed and validated predominantly in Western populations, with an underrepresentation of Asian patients. In this study, we explore the prognostic value of CanAssist Breast (CAB), a proteomic prognostic test, in optimal treatment management of patients aged ≤50 years.

**Methods:** This study includes a previously published retrospective cohort. The performance of CAB was evaluated using Kaplan-Meier analysis, with 5-year Distant recurrence-free interval (DRFI) from diagnosis as the endpoint; the study also used multivariate analysis to evaluate the independent prognostic value of CAB.

**Results:** In the retrospective cohort, CAB identified 70% as low-risk (LR) and 30% as high-risk (HR) with DRFI of 93.1% (P<0.0001); further classification showed 64% LR and 36% HR in the Asian and 75% LR and 25% HR in the Caucasian subgroup. In patients with N0 disease, CAB identified 85% as LR and 15% as HR. In N+ patients, CAB identified 49% as LR. All CAB LR patients have an acceptable DRFI of >90% at 5 years from diagnosis.

**Conclusions:** Based on the results presented, CAB adds prognostic value for patients ≤50 years and can be used as a treatment guidance tool for these patients.

## Introduction

Breast cancer (BC), a significant global health concern, is steadily increasing, with a particular rise among women under 50.^1^ This young demographic is significant because it is often associated with a more aggressive form of the disease, and patients have a long life ahead.^2^

In young women, BC management poses unusual challenges. This is not only because of the aggressive biology of the disease and poor prognostic features, but also because of the challenging treatment decisions for clinicians, keeping in mind the fertility, body image, and stress over life disruptions like work, relationships, and identity issues related to young women.^3,4^ Although typically associated with favourable outcomes in older patients, Hormone-positive (HR+), HER2-negative (HER2-) BC exhibits distinct behaviour in younger patients, demonstrating intrinsically more aggressive tumor biology, leading to higher recurrence and mortality rates.^5,6^

The reliance on standard clinical parameters and biomarkers [TNM status, Estrogen Receptor (ER), Progesterone Receptor (PR), HER2] routinely used to guide treatment in older HR+/HER2-patients appears limited in young patients.^5^ Many studies show age as an independent prognostic risk factor^7,8^ but some argue otherwise.^9^ Young women frequently undergo intensive (beyond surgery and radiation) chemotherapy, endocrine therapy, and targeted biological therapies, each of which can lead to substantial side effects and adversely affect quality of life. Furthermore, in some cases, young women may be subjected to overtreatment if prognostic assessment is only based on their age, adding to the challenges of managing diagnosis and treatment.^9^

To address these challenges, prognostic tests that incorporate tumor biology into prognostic assessments have been developed and are widely commercially available.^10,11^ Most contemporary clinical guidelines recommend using validated prognostic assays to guide adjuvant therapy decisions in early HR+ BC, including in younger patients, where traditional clinicopathologic features alone may be insufficient to inform treatment choices. ^12,13^

Among the prognostic tests available today, CanAssist Breast (CAB) is the only proteomic-based test that uses a machine-learning algorithm to predict the risk of distant recurrence. CAB combines the expression of five protein biomarkers (CD44, ABCC4, ABCC11, N-cadherin, and Pan-cadherin) involved in cancer metastasis and drug resistance measured by immunohistochemistry (IHC), and three clinical parameters (tumor size, grade, and lymph node status) to segregate patients as “Low-Risk” (LR) or “High-Risk” (HR) for distant recurrence within five years from diagnosis. Multiple retrospective validation studies across India, the USA, and Europe (Spain, Germany, Austria, Italy) have demonstrated the robust performance of CAB in predicting recurrences over 5 years.^14–18^ Additionally, the ability of CAB to predict 10-year recurrence risk has been demonstrated in the prospective, randomised, completed TEAM trial in the Netherlands.^19^

Since mid-2016, CAB has been in clinical use and utilised by 10,000+ patients from India, Turkey, the UAE, Sri Lanka, Bangladesh, and Iran to plan treatment. In routine clinical settings, CAB has demonstrated its utility in guiding clinicians in tailoring chemotherapy decisions.^20,21^ Importantly, CAB has been included in the guidelines of the Asian Geriatric Oncology Society (AGOS), Indian Society of Medical and Paediatric Oncology (ISMPO)-Early Breast Cancer management, ISMPO-Breast Cancer in Young Guidelines, and Association of Breast Surgeons of India (ABSI).^13,22–24^

The current study aims to evaluate the prognostic value of CAB in young patients (≤50 years) through data analysis of a retrospective cohort.

## Methods

### Data curation and Study design

This study includes a cohort of young patients (≤50 years) with HR+, HER2-early breast cancer (EBC) (n=833) from previously published retrospective studies that validated CAB in patients from India, the USA, and five European countries (Austria, Germany, Spain, Italy, and the Netherlands).^14–19^

### Patient inclusion, follow-up, and ethics approvals

Data analysis of previously published retrospective studies was performed in an anonymized manner in compliance with standard ethical guidelines and approved by relevant ethics committees, IRBs, or biobank approvals, as detailed in the original publications.^14–19^

### CAB test performance

IHC staining for five CAB biomarkers and grading were performed as described earlier^25,26^, generating an LR (1 to 15.5) or HR (15.6 to 100) CAB risk for distant recurrence.

### Statistical analyses

Distant recurrence-free Interval (DRFI) at 5 years after breast cancer diagnosis was estimated using Kaplan-Meier (KM) survival curves (GraphPad Prism 10). The P-value (Log-rank test) was used to assess the association between DRFI across the two risk groups; P-value <0.05 was considered statistically significant. The multivariate analysis was performed using MedCalc software (22.032–64-bit).

## Results

### Description of the retrospective ≤ 50 years cohort (n=833)

The retrospective cohort in this study comprises 833 patients who were aged 50 or younger. The majority (51%) of the patients had a T2 tumor. Interestingly, the cohort included 57% node-negative (N0) and 43% node-positive (N+) patients. Grade 2 (G2) was the most common (57%), followed by Grade 3 (G3) (33%) and Grade 1 (G1) (11%). CAB stratified 70% of patients as having a LR, while the remaining 30% were stratified as having a HR of distant recurrence (Table 1).

**Table 1:** Patient demographics and CAB-based risk stratification in the retrospective cohort.

| **Clinicopathological Features** | **Subgroups** | **No. of Patients (n)** | **Percentage of Patients (%)** |
| --- | --- | --- | --- |
|  | **Total** | 833 | 100 |
| **Tumor size** | **T1** | 365 | 44 |
|  | **T2** | 428 | 51 |
|  | **T3** | 40 | 5 |
| **Node status** | **N0** | 478 | 57 |
|  | **N+** | 355 | 43 |
| **Histological Grade** | **G1** | 88 | 11 |
|  | **G2** | 474 | 57 |
|  | **G3** | 271 | 33 |
| **Treatment Given** | **Chemotherapy Treated** | 596 | 72 |
|  | **ET Alone** | 237 | 28 |
| **CAB Risk Category** | **Low Risk** | 581 | 70 |
|  | **High Risk** | 252 | 30 |

### CAB risk stratification based on Ki-67 expression in ≤50 Years

Among the 833-patient cohort, Ki-67 information was available for 67% of patients (n=599). According to the latest guidelines from the International Ki-67 Working Group (IKWG), patients were categorized as low Ki-67 (≤5%), intermediate Ki-67 (6-29%), or high Ki-67 (≥30%) based on Ki-67 expression. We observed that 44% of the cohort had low Ki-67 expression, followed closely by 39% with intermediate expression, and only 17% with high expression (Figure 1a).

**Figure 1:**
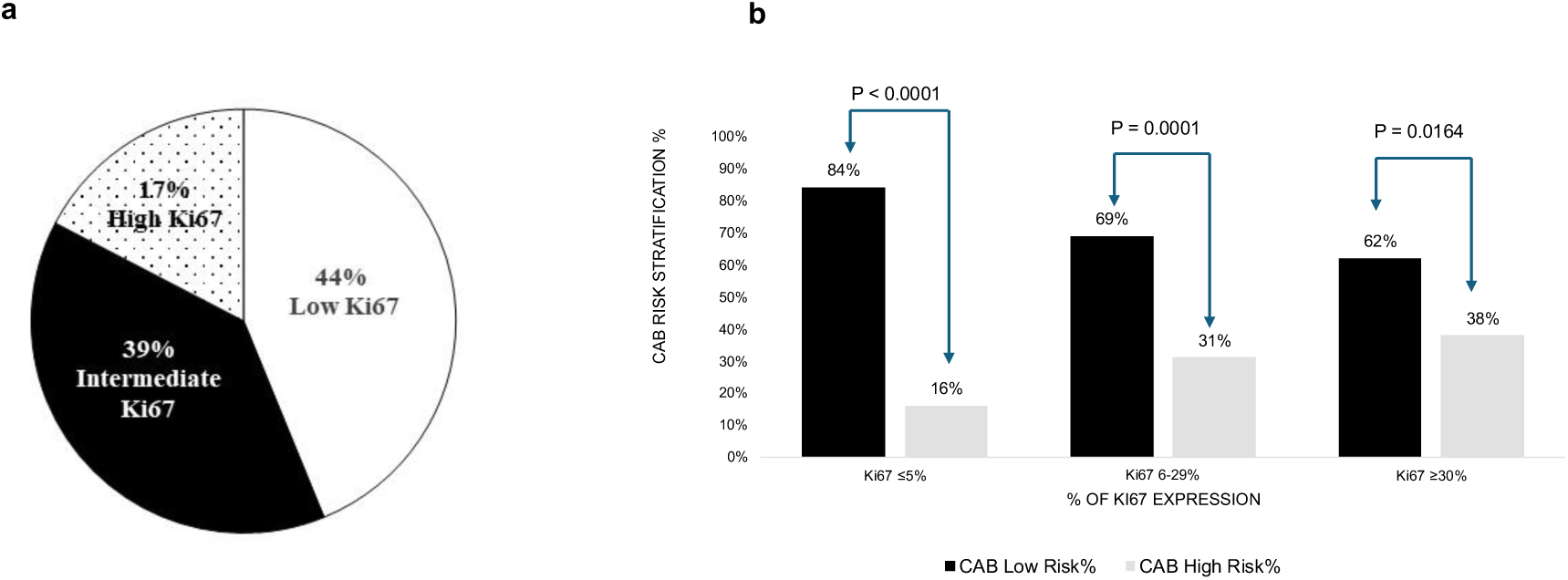
Use of CAB in different Ki67 subgroups. (a) Pie chart showing the percentage of patients in different Ki67 subgroups (classified according to the IKWG guidelines). (b) Graph displaying CAB risk stratification in Low Ki67 (≤5%), Intermediate Ki67 (6-29%), and High Ki67 (≥30%) subgroups. The chi-squared test was used to calculate P-values by using MedCalc software.

CAB significantly stratified patients with low Ki-67 expression (84:16, P < 0.0001) and those with high Ki-67 expression (62:38, P = 0.0164) into LR: HR groups. Notably, CAB stratified 16% of patients as HR in the low-Ki67 subgroup and 62% as LR in the high-Ki67 subgroup (Figure 1b). Interestingly, in the intermediate Ki-67 subgroup, CAB provided clearer risk stratification, with significant LR: HR proportions of 69:31 (*P*=0.0001).

### Subgroup-Specific Hazard Ratios for Recurrence According to CAB Risk

The subgroup analysis of clinical features based on CAB risk stratification showed promising hazard ratios. In Figure 2, the Forest plot demonstrated consistent prognostic stratification by CAB across clinically relevant subgroups. Patients classified as HR by CAB had increased hazards compared to the CAB LR patients in the overall cohort and in clinical subgroups such as age, grade, and tumor size.

**Figure 2:**
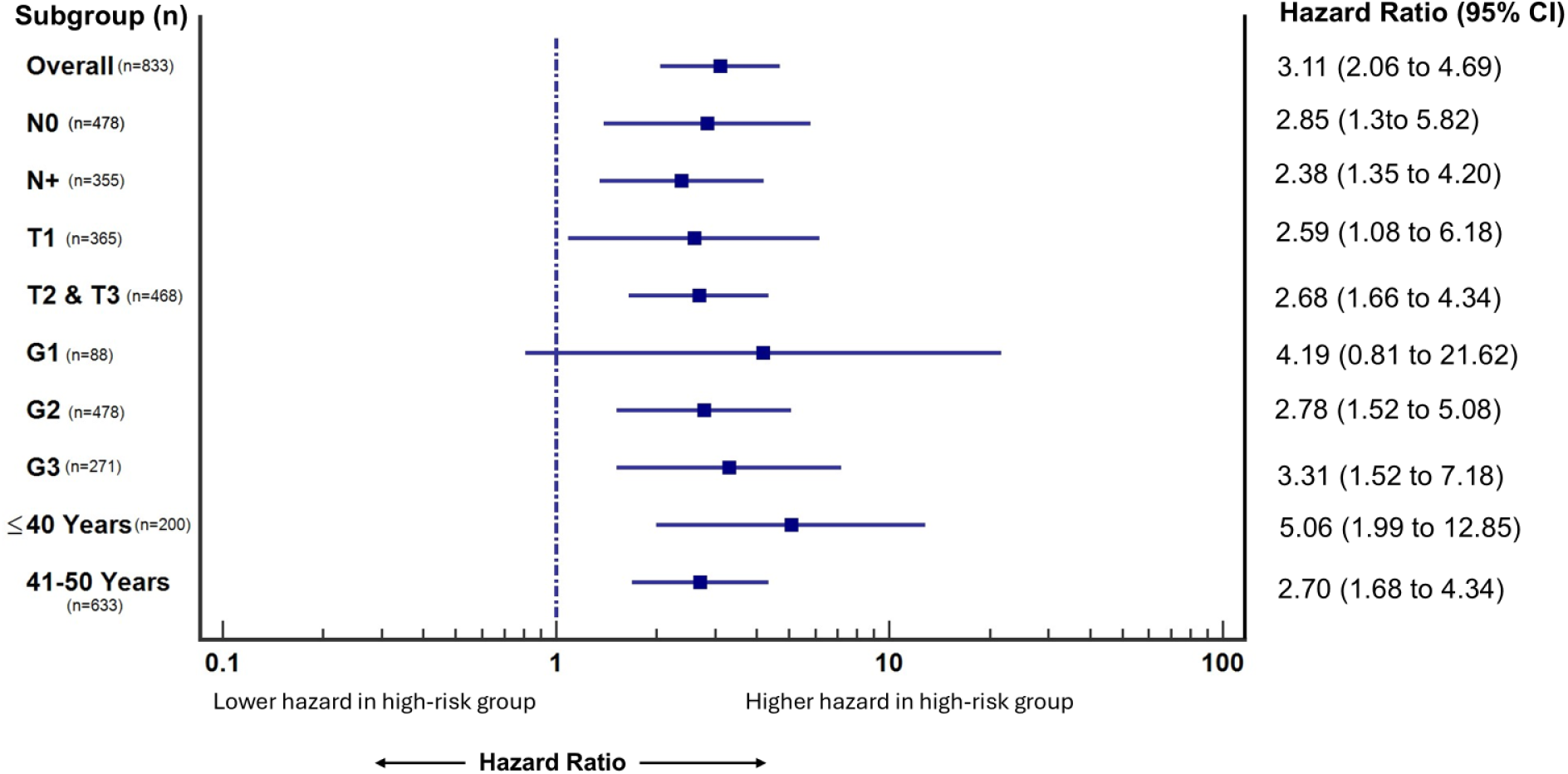
Forest plot showcasing CAB risk stratification. The Forest plot shows consistent prognostic stratification by CAB across clinically relevant subgroups. *CI, confidence interval*

### CAB demonstrates independent prognostic value

As shown in Table 2, a multivariate analysis was performed to assess the ability of CAB to serve as an independent prognostic indicator. This analysis included the CAB risk categories along with clinical parameters such as grade, node status, and tumor size. Among these, we observed that tumor size (HR = 2.0, 95% CI = 1.2322 to 3.2807; P = 0.0052) and CAB showed a significant association with outcomes. However, CAB showed greater prognostic significance, with a hazard ratio (HR) of 2.27 (95% CI, 1.4194 to 3.6574; P = 0.0006).

**Table 2:** Multivariate analysis of the young patient cohort for distant recurrence.

| Covariate | Hazard ratio | P-value | 95% CI |
| --- | --- | --- | --- |
| **Grade: G1, G2 + G3** | 1.0267 | 0.9075 | 0.6587 to 1.6001 |
| **Node status: N0, N+** | 1.5057 | 0.0791 | 0.9536 to 2.3776 |
| **Tumor size: ≤T1, >T1** | 2.0106 | 0.0052 | 1.2322 to 3.2807 |
| **CAB risk score: low, high** | 2.2785 | 0.0006 | 1.4194 to 3.6574 |

*CI: confidence interval.*

### Survival analysis of young patients in the retrospective cohort

KM survival analysis at 5 years from diagnosis demonstrated significant risk stratification by CAB in patients aged ≤50 years (n=833) and in a subgroup of patients (n=237) who had received endocrine therapy (ET-alone) (Figure 3a, 3b). In Figure 3a, 70% of these patients were stratified as LR by CAB and had an acceptable DRFI of 93.1% irrespective of chemotherapy use. In contrast, ∼30% of patients from the HR category had a significantly worse DRFI of 79.7% (P < 0.0001); this is comparable to the ET-alone cohort shown in Figure 3b (P = 0.0119). Analysis in the Asian population (n=419) showed that CAB identified 64% of patients as LR and 36% as HR, with an LR DRFI of 94.1%. On the other hand, CAB stratified 75% of patients as LR and 25% as HR in the Caucasian population (n=414) with an LR DRFI of 92.2% (Figure 3c, 3d).

**Figure 3:**
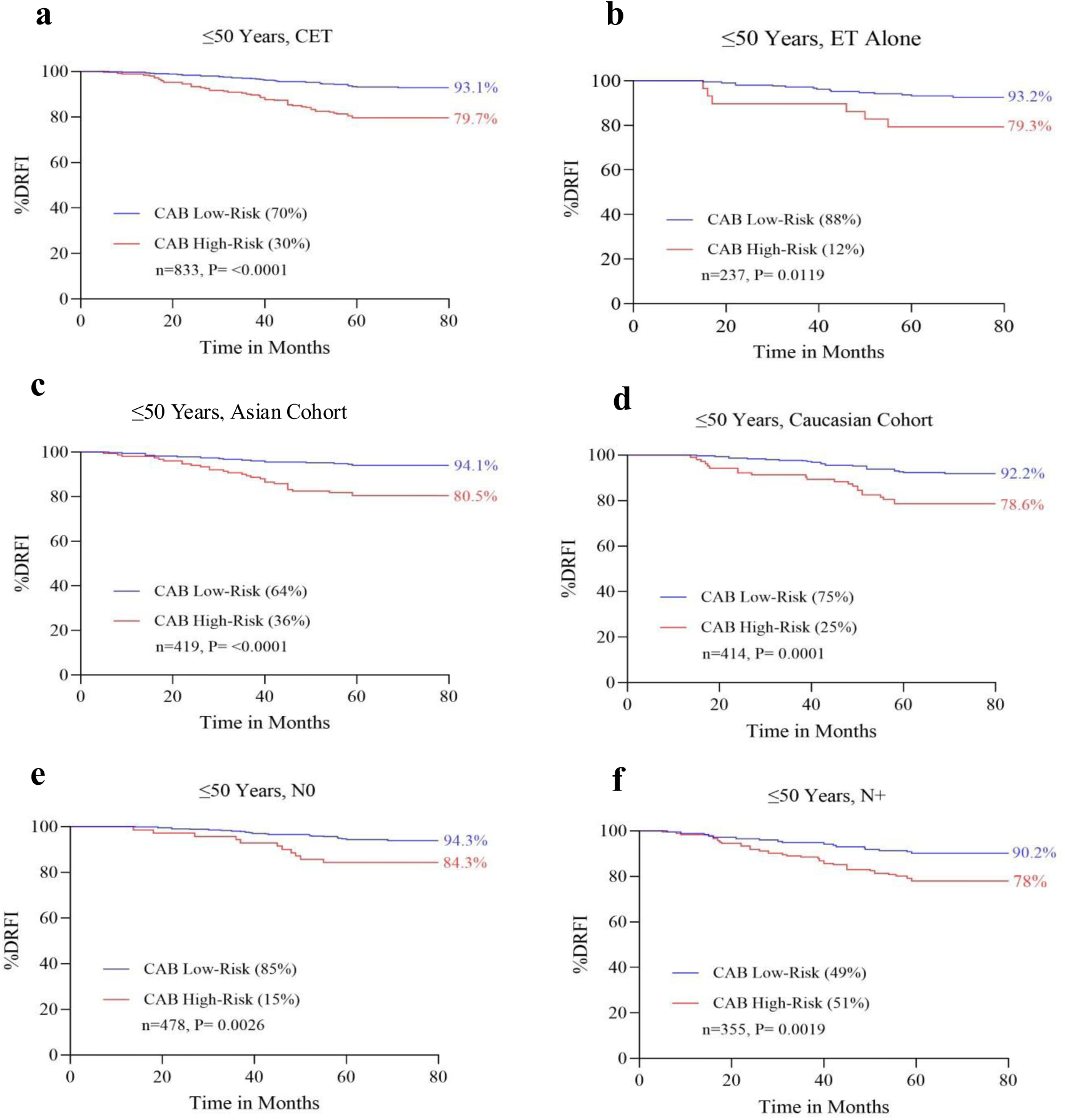
Performance of CAB in a retrospective young patient cohort. Kaplan-Meier survival curves for DRFI at five years (60 months) from diagnosis. (a) ≤50 years patients’ total cohort (CET treated) (b) subset treated with ET-alone (c) ≤50 years Asian patients’ subgroup (d) ≤50 years Caucasian patients’ subgroup (e) ≤50 years patients with N0 tumors (f), and ≤50 years patients with N+ tumors. CET: All patients, ET: Endocrine therapy alone, DRFI: Distant Recurrence-Free Interval

In patients ≤ 50 years, N0 patients (n=478), an impressive and significant DRFI of 94.3% was observed in 85% of CAB LR patients, and a DRFI of 84.3% in CAB HR patients (P = 0.0026) (Figure 3e). We further analysed N+ patients (n=355) and noted an acceptable DRFI of 90.2% in 49% of CAB LR patients, and an inferior DRFI of 78% in CAB HR patients (P = 0.0019) (Figure 3f).

## Discussion

In this study, we evaluated the prognostic value of CAB in young (≤50 years) HR+, HER2-EBC patients. Recent studies reviewing treatment strategies in young patients have shown that risk stratification based on clinicopathological features alone can lead to over- or undertreatment.^27,28^ The majority of young patients are keen to preserve their fertility in the immediate future and therefore prefer to avoid or postpone chemotherapy.^29^ Therefore, prognostic tests help bridge the gap by making tumor biology the primary factor in risk assessment and subsequent treatment decisions.

Western prognostic tests, such as Oncotype DX and MammaPrint, have trials such as TAILORx, RxPONDER, and MINDACT that report chemotherapy benefit in young women, but young Asian women are underrepresented in these trials.^30–32^ Asian countries recorded the highest number of BC cases globally, with a substantial proportion occurring in women under 50 years.^33^ Presentation of EBC in Asian patients is distinct. Asian women are diagnosed at a younger median age than their Western counterparts, and Asian patients have a different underlying tumor biology.^24^ Therefore, it is necessary to have a prognostic tool validated specifically in the Asian population. To address this, we explored the potential of CAB - a test developed on Asian patients - in this demographic. Previous retrospective studies have established concordance between CAB and Western tests such as Oncotype DX and MammaPrint, with a low-risk concordance of 83%. ^17^

In the current retrospective study cohort, CAB stratified the majority of the cohort as LR, with an observed five-year DRFI of 93.1%, suggesting improved survival outcomes compared with HR patients. We further demonstrate the utility of CAB in the Asian and Caucasian subgroups. CAB stratified 11% more Asian patients as HR compared to Caucasian patients. This observation aligns with the inherent differences in tumor biology, age at diagnosis, and disease presentation, which have been previously observed among Asian and Caucasian populations. ^24^Despite differences in risk categorization, DRFI across both subgroups is comparable, with 94.1 versus 92.2 in LR and 80.5 versus 78.6 in HR, respectively. This indicates that the biological risk measured by CAB is clinically meaningful and reproducible across Asian and Caucasian populations.

Young node-positive patients (clinically high-risk) would traditionally be treated with chemotherapy. In the retrospective cohort analysis, CAB stratified ∼50% of patients in this subgroup as LR of recurrence with an acceptable DRFI of 90.2%, which would prevent over-treatment for these patients. Interestingly, in young node-negative patients, CAB stratified 85% of patients for LR of recurrence with excellent DRFI (94.3%).

About 7% of young women under 40 in developed countries and 25% in developing countries are diagnosed with BC.^34^ Additionally, there is a very worrying trend of an increase in breast cancer in young women. These young women are generally associated with poor prognosis and a higher risk of recurrence due to tumor biology. A previous study by Azim et al. discusses several mutations that contribute to the aggressive nature of BC in young women and highlights the use of prognostic tests to identify high-risk mutations and distinct gene expression patterns, guiding personalized treatment strategies.^35^ Studies have shown that adjuvant ovarian function suppression (OFS) in combination with ET significantly improved recurrence-free survival in premenopausal patients with BC.^36,37^ In our previously published analysis of patients ≤40 years, CAB stratified 62% of patients as LR, with a DRFI of 95%, and also identified 38% of patients as HR, who performed considerably worse, with a 78% DRFI. ^38^ It is important to remember that all of these patients were treated with OFS drugs, indicating that additional therapies (beyond OFS) may be selectively given to young CAB HR patients to improve their outcomes.

A recently published real-world data also showcased improved adherence to treatment recommendations among young patients^20^ and the inclusion of the CAB test in the ISMPO guidelines for Breast Cancer in Young patients, underscores its role in guiding treatment decisions in this patient population^13^. Taken together, these findings show that CAB could help a majority of young LR patients avoid chemotherapy and demonstrate its potential as a comparable prognostic test with Western tests available in the market.

The notable strength of the study is the equal representation of Asian and Caucasian populations in the overall cohort. We also acknowledge our limitations; its retrospective design may introduce selection bias.

## Conclusion

CAB provides valuable prognostic information for patients with HR+ / HER2-EBC aged ≤50 years, supported by robust data. These findings further suggest that using CAB could improve risk assessments in these ≤50-year-old at-risk patients and help clinicians refine treatment strategies for them.

## Acknowledgements

We thank Mr. Manjunatha G, Mrs. Prathima R, Mr. Harishkumar N, and Mr. Dinesh Babu P from OncoStem Diagnostics, Bangalore, for their help with IHC experiments and histopathological work.

## Declaration of generative AI and AI-assisted technologies in the writing process

During the preparation of this work, the authors used ChatGPT (OpenAI) and Grammarly to improve language and readability. After using these tools/services, the authors reviewed and edited the content as needed and take full responsibility for the publication’s content.

## Data Availability

The data generated during and/or analysed during the current study are available from the corresponding author upon reasonable request.

## CRediT authorship contribution statement

**Manvi Sunder:** Writing-Original Draft, Data curation, Formal analysis. **Tejal D. Durgekar:** Writing-review & editing, Data curation, Formal analysis**. Shyamili Goutham:** Data curation, Formal analysis. **Badada Ananthamurthy Savitha:** Data curation. **Payal Shrivastava:** Data curation. **Naveen Krishnamoorthy:** Data curation. **Deepti K Shivashimpi:** Data curation. **Koushik Anand S J:** Data curation. **Anjali Raghuram:** Data curation. **Manjiri M. Bakre:** Conceptualization, Validation, Supervision, Formal Analysis, Writing-review & editing.

